# Psilocybin alters brain activity related to sensory and cognitive processing in a time-dependent manner

**DOI:** 10.1101/2024.09.09.24313316

**Authors:** Marek Nikolic, Pedro Mediano, Tom Froese, David Reydellet, Tomas Palenicek

## Abstract

Psilocybin is a classic psychedelic and a novel treatment for mood disorders. Psilocybin induces dose-dependent transient (4-6 hours) usually pleasant changes in perception, cognition, and emotion by non-selectively agonizing the 5-HT_2A_ receptors and negatively regulating serotonin reuptake, and long-term positive antidepressant effect on mood and well-being. Long-term effects are ascribed to the psychological quality of the acute experience, increase in synaptodensity and temporary (1-week) down-regulation of 5-HT_2A_ receptors. Electroencephalography, a non-invasive neuroimaging tool, can track the acute effects of psilocybin; these include the suppression of alpha activity, decreased global connectivity, and increased brain entropy (i.e. brain signal diversity) in eyes-closed resting-state. However, few studies investigated how these modalities are affected together through the psychedelic experience. The current research aimed to evaluate the psilocybin intoxication temporal EEG profile. 20 healthy individuals (10 women) underwent oral administration of psilocybin (0.26 *mg/kg* ) as part of a placebo-controlled cross-over study, resting-state 5-minute eyes closed EEG was obtained at baseline and 1, 1.5, 3, 6, and 24 hours after psilocybin administration. Absolute power, relative power spectral density (PSD), power envelope global functional connectivity (GFC), Lempel-Ziv complexity (LZ), and a Complexity via State-Space Entropy Rate (CSER) were obtained together with measures of subjective intensity of experience. Absolute power decreased in alpha and beta band, but increased in delta and gamma frequencies. 24h later was observed a broadband decrease. The PSD showed a decrease in alpha occipitally between 1 and 3 hours and a decrease in beta frontally at 3 hours, but power spectra distribution stayed the same 24h later. The GFC showed decrease acutely at 1, 1.5, and 3 hours in the alpha band. LZ and showed an increase at 1 and 1.5 hours. Decomposition of CSER into functional bands shows a decrease in alpha band but increase over higher frequencies. Further, complexity over a source space showed opposing changes in the Default Mode Network (DMN) and visual network between conditions, suggesting a relationship between signal complexity, stimulus integration, and perception of self. In an exploratory attempt, we found that a change in gamma GFC in DMN correlates with oceanic boundlessness. Psychological effects of psilocybin may be wrapped in personal interpretations and history unrelated to underlying neurobiological changes, but changes to perception of self may be bound to perceived loss of boundary based on whole brain synchrony with the DMN in higher frequency bands.

## 1 Introduction

Psilocybin is studied as a classic psychedelic and a novel antidepressant which induces potent changes to consciousness, alongside with lysergic acid diethylamide (LSD) and dimenthil triptamine (DMT) and other related chemicals of similar effects. Psilocybin’s active metabolite psilocin is a non-selective serotonin 5-HT receptor agonist and SERT inhibitor (Dinis-Oliveira, 2017; Rickli et al., 2016). The antidepressant effect of a psychedelic dose of psilocybin has a rapid onset and sustained duration (R. Carhart-Harris et al., 2021; Davis et al., 2021; Griffiths et al., 2016; Yu et al., 2022). Evidence from animal research suggests that these effects relate to lasting increased synaptogenesis and transient (<week) downregulation of 5-HT_2A_ receptors (Raval et al., 2021) in the hippocampus and prefrontal cortex. The antidepressant effect was previously associated with the magnitude of alterations to cognition, emotion, and perception (Roseman et al., 2018) induced by agonisation specifically of the 5-HT_2A_-Gq/11 receptors (Wallach et al., 2023), however, complete remission in a treatment-resistant depression patient after single-dose of psilocybin and pretreatment with a 5-HT_2A_ occupant trazadone was reported (Rosenblat et al., 2023), suggesting that psychedelic experience may be a useful marker of intensity of the experience, but not necessary for the long-term effects. The acute experience is accompanied by an increase in body temperature, heart rate, and blood pressure with return to normal within 12 hours (Holze et al., 2022). Early evidence suggests that increase in blood pressure and heart rate may be a marker of antidepressant response (PhD Thesis, Andrashko).

Subjective psychedelic experience is generally characterised by intense changes in perception, emotion, and cognition, changes usually perceived as positive (Studerus et al., 2011). These changes are occasionally accompanied by feelings of dysphoria and panic in high doses, and show potential to be resolved towards positive outcomes when occurring in supportive setting (Kluckova et al., 2024; Studerus et al., 2011). Clinical use of psilocybin does not lead to subsequent drug abuse or increased incidence of mental health problems (Krebs & Johansen, 2013; Studerus et al., 2011). Intensity of psychedelic experience is captured by an Altered State of Consciousness questionnaire (ASC; Dittrich, 1998). The main axes of the experience are oceaninc boundlessness (OBN), anxious ego-dissolution (AED), and visionary restructuralization (VIR). Generally OBN captures the mystical nature of the experience, VIR captures the hallucinatory part, whereas AED captures the aspects of a ’bad trip’. The limitation of the ASC is that it provides a narrow retrospective narrative of the experience void of personal process (Preller & Vollenweider, 2018).

Electroencephalography (EEG) captures the electrical activity generated by large groups of neurons, offers a convenient, cost-effective, user-friendly, noninvasive method with high temporal resolution suitable for capturing changes in brain activity in humans. Bravermanová et al., 2018 show that acute intoxication with psilocybin affected early perceptual (p100) and higher cognitive (p300) processes and that the size of this effect correlated with the individual intensity of the experience but that pre-attentive cognition was not affected. The study of a resting-state (task-free activity), usually described in terms of neural synchrony (power and functional connectivity) and predictability (signal complexity) shows that the psychedelic state is hallmarked by a suppression of alpha (8-12Hz) power spectral density acutely (in psychedelics, in general, Pallavicini et al., 2019, in psilocybin accompanied by a theta desynchronization Ort et al., 2023), an increase in functional connectivity post intoxication (in the Default Mode Network (DMN), Kuburi et al., 2022), and an increase in brain signal diversity (Lempel Ziv complexity LZc; Lempel and Ziv, 1976, LSD, Mediano et al., 2020, DMT Timmermann et al., 2019, psilocybin M. M. Schartner et al., 2017). Psilocybin modulates perceptual and higher-order activity to be more diverse and unpredictable while preserving complex causal functioning (Bravermanová et al., 2018; Ort et al., 2023).

Capturing progressive changes induced by the onset and duration of the psychedelic experience showed power spectral density desynchronization (delta 0-4Hz, alpha 8-13Hz, and gamma 30-45Hz) dependent on plasma levels, but changes in complexity (LZc) peaked after the intensity of experience started to recede (DMT; Timmermann et al., 2023). Sensitivity of LZc to changes lasting beyond the acute intoxication may be explained as relating to changes in thought pattern often reported by participants after a psychedelic experience (Mason et al., 2019). LZc is dependent on level of interaction with environment; increase in LZc was the largest in an eyes-closed resting-state condition compared to music condition, eyes open, and TV watching; activities which also increase brain entropy, but this increase was not as pronounced under LSD when compared with placebo (Mediano et al., 2024). Considering the visual nature of psychedelic experience especially in eyes closed condition, together with that visual stimulation increases Lempel-Ziv diversity (Orłowski & Bola, 2023), it may be that changes induced by psychedelics turn trivial stimulation into vivid experiences, an effect pronounced particularly in the absence of any stimuli (Kometer & Vollenweider, 2018; Wießner et al., 2021). Complexity via State-Space Entropy Rate (CSER) is a novel measure analogical to LZc, which allows the study of brain signal diversity across functional bands, thus crossing the bridge between LZc, and classical power analysis. Mediano et al., 2023 show that the effect of psychedelics on brain entropy is mostly in the fast oscillating gamma band (> 25Hz), associated with higher order activity.

The continual changes in brain activity have insofar not been tested under psilocybin using EEG. We expect to see changes in absolute power, relative power spectral density, mainly in the alpha (8-12Hz). We expect to see changes in brain signal complexity extended beyond the acute effects, and that these changes will be mostly in the gamma frequency. We expect a global desynchronisation. We expect to find an increase in LZc. We expect the magnitude of these changes to correlate with the intensity of the experience measured by ASC, particularly with changes in the DMN. Acute effects should be accompanied by a transient changes to heart rate but not heart rate variability. An exploratory attempt will be made at relating EEG measures to the intensity of the experience, and at correlating the GFC and CSER.

## 2 Methods

### 2.1 Study approval

The study was approved by the Ethical Committee of the National Institute of Mental Health (NIMH–CZ), by the State Institute for Drug Control, and as a clinical trial registered under the EudraCT No. 2012-004579–37.

### 2.2 Experimental design

In a double-blind, placebo-controlled crossover design, all participants took part in active and placebo sessions, which were a minimum of 28 days apart (thought to remove any subacute effects or afterglow of psilocybin Aday et al., 2020; Evens et al., 2023; Studerus et al., 2011, an average of 67.55 days apart, SD = 69.53). This experiment had two arms: EEG and fMRI (see fig. 2.2). The current experiment was part of the EEG arm and was not affected by the fMRI arm that followed. As the EEG arm always preceded the fMRI arm, participants were unblinded to which arm they were participating in.

**Figure 1:**
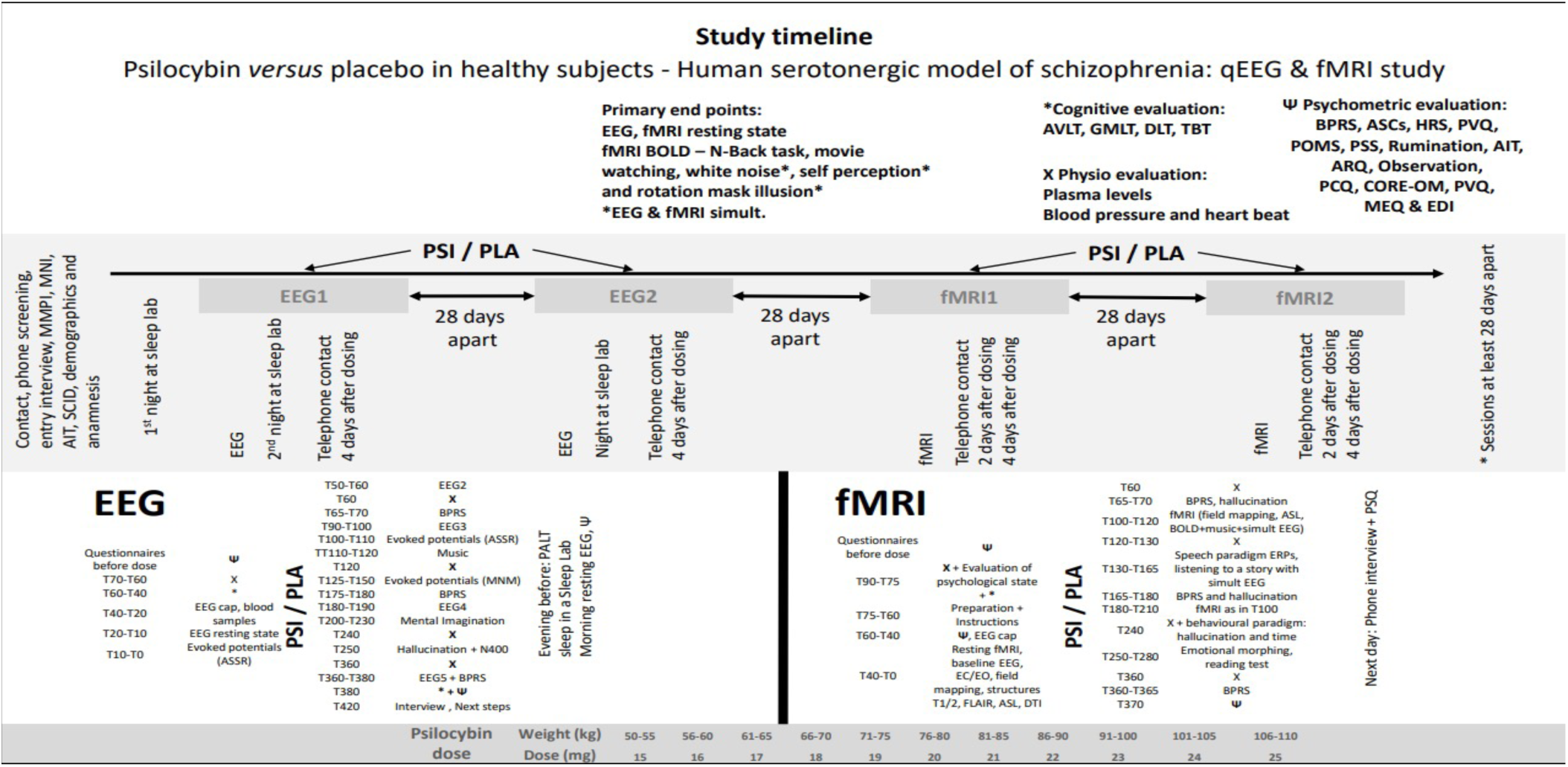
Study timeline: current study was a part of a larger clinical trial with many research caveats. From Kluckova et al., 2024.

**Figure 2:**
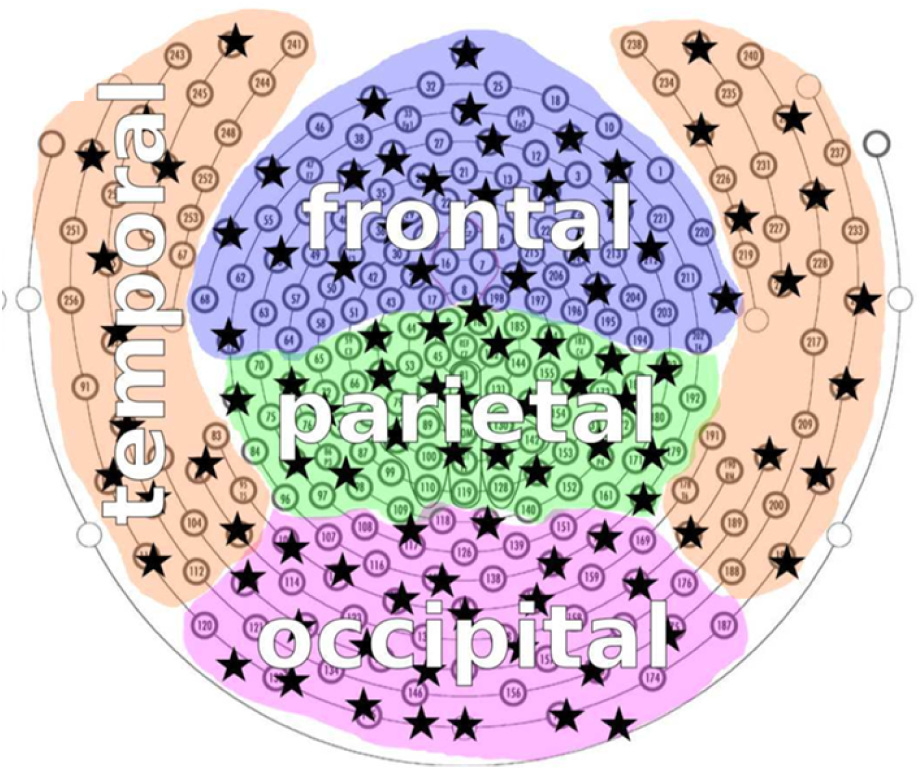
Electrodes divided by brain regions. Left and right hemisphere were divided at the central line. Image from M. Schartner et al., 2015.

The order of drug administration, gender, and order of drug/placebo administration was randomized in 5 balanced blocks of 4 participants. Psilocybin and placebo capsules were prepared by an independent, unblinded pharmacy. Each session began with a short examination of participants, assuring the exclusion of acute contraindications for participation. This included somatic examination, questions about a current life situation, blood pressure, and heart rate, and a negative result of alcohol breathalyzer and urine rapid drug tests for main drug classes. The session was held in a living room-like decorated experimental room inside a Faraday cage where subjects spent most of the experiment. During the session, they were always accompanied by a gender-balanced team of two sitters, one of whom was always a psychiatrist. The study nurse was also present in a neighboring room throughout the session.

Participants slept at the clinic for habituation. On the morning of the experimental days (around 9 am), psilocybin was administered orally on an empty stomach with 200 ml of water in 1 and 5 mg capsules in a dosage adjusted to body weight (1 mg per 5 kg, range 15–22 mg, mean = 18.35 mg, SD = 2.21 mg). Psilocybin-induced psychedelic effects last approximately 6 hours. A standardized music playlist was played throughout most of the session and was paused for the EEG recording. Approximately 9 hours after the dosing, participants went to sleep in a sleep lab.

### 2.3 Participants

Twenty healthy volunteers (10 M/10F, mean age = 36, SD = 8.1, range 28 - 53) took part in the study as part of a larger clinical trial registered under *EudraCT No. 2012-004579-37* (major inclusion criteria are in appendix A.1, for full recruitment, screening, and procedure details see Viktorin et al., 2022). Subjects were recruited from November 2017 to July 2018 via a peer-to-peer method and pre-screened via phone or email interview to exclude those who did not meet the major inclusion criteria. Those that met the major screening criteria were invited to an introductory meeting. During a two-hour-long interview, participants were informed in detail about the nature of the study, the drug effects, and safety. They were free to ask questions related to the experiments or psilocybin. All participants provided written informed consent before they entered the study. Subjects were screened by the Minnesota Multiphasic Personality Inventory (MMPI-2, Butcher et al., 1991) and Mini-International Neuropsychiatric Interview (MINI, Sheehan et al., 1998) for any significant psychopathology. The study design corresponded to the Guidelines for Safety in Human Hallucinogen Research (Johnson et al., 2008). Participants were asked to abstain from drug use during the period between the interview and the experiment, alcohol use leading to drunkenness (1 week), coffee and food (in the morning), and tobacco (2 h) before the experiment. One participant was excluded due to excessive daytime sleepiness (recognized during the placebo EEG session). To avoid interactions with the menstrual cycle, women were not dosed during their menses (see Dudysova et al., 2020 for details).

### 2.4 EEG aquisition

20 participants were included in the study. One participant did not arrive to the second session. 4 recordings had to be excluded for non-systematic mistakes including detached reference electrode (2), a mistake during saving the recording (1), and one missing file. Data were excluded element-wise, resulting in 230 data entries included in the analysis.

### 2.5 Psilocybin dose

Psilocybin was manufactured according to good manufacturing practice standards from THC-Pharm GmbH, Frankfurt, Germany. Gelatine capsules containing 1 and 5 mg of psilocybin homogenized with *Trittici amylum* were prepared in the pharmacy of the Institute for Clinical and Experimental Medicine in Prague, Czech Republic. The dose was set to be approximately 0.26 *mg/kg*, an intermediate psychedelic dose (Nichols, 2016; Tyls et al., 2016), and was adjusted by 1mg per each 5kg of body weight. The drug was administered orally in an adjusted number of capsules and swallowed with drinking 200 mL of water.

### 2.6 Acute psychedelic state

The Brief Psychiatric Rating Scale (BPRS), commonly used for a brief evaluation of psychiatric symptoms by a clinician, informs about the absence, presence, and severity of symptoms that include anxiety, depression, hostility, grandiosity, hallucinations, and unusual thought content (for full details, see Overall and Gorham, 1962). In the context of our trial it is used to rate the intensity of the clinically relevant psychological impact of psilocybin across the session at baseline, 1-, 3-, and 6-hours after (Figure 4). Participants were evaluated by a psychiatrist-rater blinded to the treatment condition.

The Altered State of Consciousness Questionnaire (Dittrich, 1998) is used to retrospectively evaluate the intensity of altered state of consciousness along three axis: Oceanic Boundlesssness (OBN), Anxious Ego Dissolution (AED), and Visionary Restructuralisation (VIR). Generally, OBN refers to the positive experiences of loss of boundaries and experience of unity, AED refers to the negative experiences usually related to anxiety and loss of control, and VIR captures extent to which the experience is unusual on sensory level. A revised version of the ASC, the 5d ASC includes psychedelic specific axes Auditory Alterations and Vigilance Reduction (for full details see Preller and Vollenweider, 2018. In the current study, the questionnaire was used in it’s original form.

### 2.7 EEG setup, recording intervals

A high-density equidistant 256-channel gel cap (HydroCelTM, Geodesic Sensor Net 256-channel used with EGI Net Amps) was applied by an EEG technician. EEG recordings took place in the EEG experimental room (Faraday cage), with a battery-powered EGI system (EGI NetAmp GES 400 with FICS box) sampled at 1000*Hz*. Each recording commenced with a measurement of impedance and electrode readjustment to minimize impedances. A 5-minute-long recording of resting state task-free eyes closed spontaneous activity in a semi-reclined or reclined position was obtained at baseline (prior to psilocybin/placebo ingestion) and post ingestion at 1-, 1.5-, 3-, 6-, and 24-hours intervals (see fig. 2.2 for a full protocol). Music was stopped during the resting-state recordings. The EEG cap was mounted at baseline, and kept through the 1-, 1.5-, 3-, and 6-hours data acquisition points. After 24-hours was data was acquired with a newly mounted cap.

#### 2.7.1 Data pre-processing

Pre-processing was performed using the BrainVision Analyzer (“BrainVision Analyzer (Version 2.2.2), Gilching, Germany: Brain Products GmbH.”, 2021) for all 230 acquired recordings. Data sampled at 1000*Hz* was band-pass filtered between 0.5 and 250*Hz* using an IIR filter, with a notch filter at 50*Hz*. Continuous data was visually inspected by an EEG expert; gross artifacts were marked as bad segments. An Independent Component Analysis was used to remove eye artifacts (horizontal and vertical eye movements) and ECG - ECG channel was included in the component calculation estimate. Sleep specialist marked epochs of sleep, drowsiness, and reduced vigilance. If more than 60% of the signal had to be removed due to artifacts or sleep, or if more than 4 independent components had to be removed, recording was removed from the analysis. Data was parceled into 4 second epochs with 1 second overlap and re-referenced to an average reference. This data was exported as continuous EEG data files and used in further analysis.

#### 2.7.2 Electrocardiogram

ECG was recorded and used to subtract ECG artifact from the EEG signal. QRS peaks were detected using an inhouse python script. Heart rate (HR) and heart rate variability (HRV) obtained from the peaks to compare between conditions across times.

#### 2.7.3 EEG Measures

##### Absolute power

was calculated using the fast Fourier transformation from the 4 second segments further split into 2 second segments to achieve 0.5Hz accuracy, using the compute_psd function with multi-taper option and frequencies of interest 1 to 100hz, included in the MNE library. The median was computed across epochs and channels. The % difference from baseline was computed. Further, baseline distribution of power within frequency bins were used to calculate Z score normalised changes from baseline for each participant. This step also serves to aid visual inspection of data as it removes the 1/frequency power distribution. Finally, Z scores were averaged using the median, and placebo condition was removed from psilocybin condition.

##### Relative power

was computed from the absolute power to assess the changes in spectra composition. Data were divided into the functional bands delta 1-4Hz, theta 4-8Hz, alpha 8-12Hz, beta 13-25Hz, and gamma 25-45Hz. Summative power in each band was divided by the total broadband energy, normalising the power to values between 0 and 1. Cluster analysis was conducted between Placebo and Psilocybin to assess the changes in EEG bands in time with permutation based correction for multiple comparisons.

##### Alpha peak

frequency with maximum energy was identified to analyse shift in alpha peak across conditions and times. To account for possible variation outside of the functional band, area from 6 to 15Hz was searched for maximum relative power value. Baseline subtracted values were then fitted to a Linear Mixed Model.

##### Functional connectivity

power envelope correlation was computed on orthogonalised time series FIR band pass filtered to EEG functional bands, computing the power envelopes using the Hilbert transform, then computing the correlation matrices using the mne_connectivity package. Orthogonalisation was done pairwise. Correlation matrix was averaged to obtain an average correlation from each electrode to all other electrodes (global functional connectivity, GFC), and GFC was averaged over the frontal, parietal, and occipital areas laterally, and over RSNs on source localised data.

##### Lempel Ziv

(LZ) signal diversity was calculated using the LZ76 algorithm. The algorithm was applied to 4-seconds segments Z-scored EEG downsampled to 250Hz data, then averaged channels, and over epochs, to avoid concatenation. LZ scores were normalised to obtain entropy rate.

##### Complexity via State Space Entropy Rate

(CSER) was calculated analogically to LZ (for methodology see Mediano et al., 2023) on Z-scored EEG time series downsampled to 250Hz, then integrated over the EEG functional bands. The benefit of such band decomposition is that the summation of the parts is equal to the broadband entropy, which would not be the case if data were filtered before the CSER calculation.

#### 2.7.4 Source Localisation

Source localisation was applied to pre-processed data using beamformer solution implemented in the Matlab Fieldtrip (Oostenveld et al., 2011). The LCMV method was used to obtain virtual sensors in the 90AAL atlas source-space (Rolls et al., 2020; Tzourio-Mazoyer et al., 2002), using an average BEM conduction model, and general channel locations for the HydroCel Geodesic 256-channel Sensor Net. Results from source decomposition were in statistical analysis averaged over 3 functional networks, Frontoparietal network (FPN), Default Mode Network (DMN), and the Visual Network (VIS) according to Oliver et al., 2019, and remaining space (Other, for reference see fig. 3).

**Figure 3:**
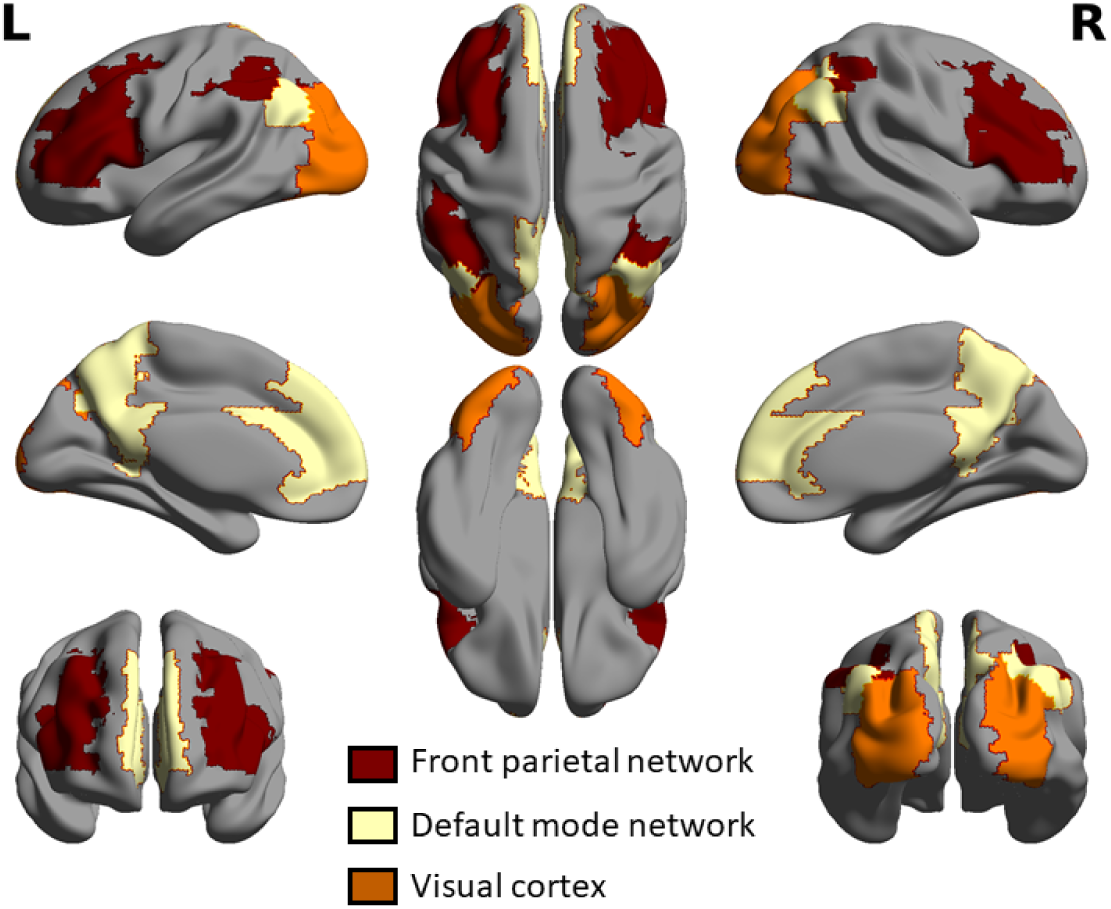
Resting state networks in the AAL90 space.

**Figure 4:**
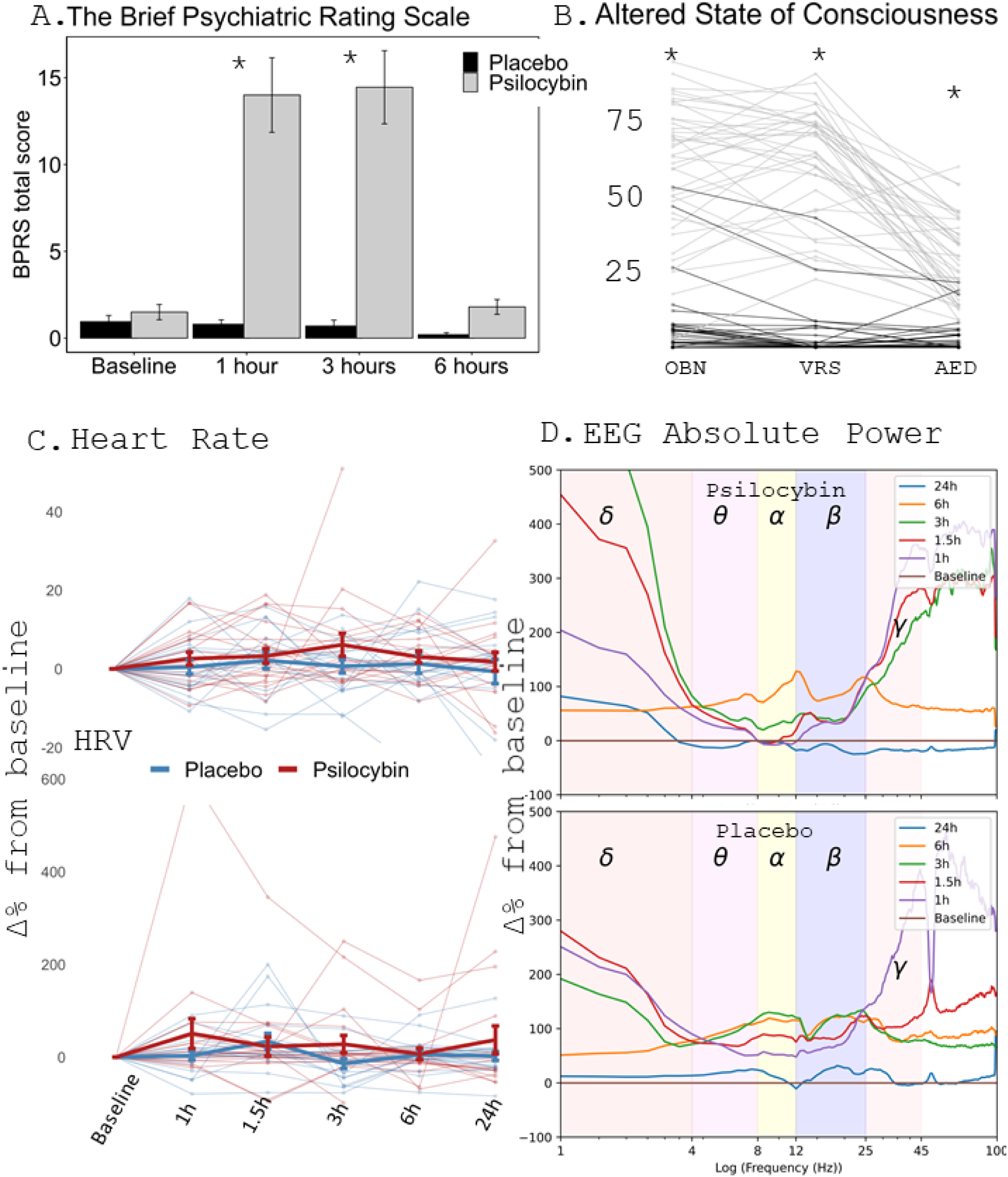
**A.** The BPRS used to evaluate mental state objectively shows significant departure from normality at 1 and 3 hours. **B.** The ASC retrospective assessment of the experience shows sig. difference between groups along the three main subscales. Note the overlap between the groups in 1̃0% of cases. **C.** shows that heart rate and heart rate variability did not differ between the groups significantly at any time point, change from baseline displayed. **D.** shows descriptive change from baseline of absolute EEG power between 1 to 100Hz. X axis is log scaled. The large differences between delta and gamma were explored further (see fig. 5.)

#### 2.7.5 Statistical analysis

Questionnaires were analysed using a t-test. Linear mixed effects models (LMM) were fit to normalised measures of EEG activity using the lme4 library for R, and the linear mixed effects models included in the statsmodels library for Python. Fixed effects of condition with two levels of psilocybin and placebo with the random effects of subject and time were included in the analysis. Analysis of variance was used to interpret results of LMM modelling. False discovery rate of 5% was maintained by adjusting all *p* values using the *p.adjust* function included in R language. Thus 5% of significant results is expected to be truly null, with the exception of the permutation based cluster analysis of relative EEG power, where missing data were removed pair-wise (for each condition across levels of time), and probability is adjusted to it’s permuted distribution.

#### 2.7.6 Exploratory analysis

In an attempt to relate different measures of the EEG signal together, we explored the changes of EEG measures in the DMN, and intensity of subjective experience measured by the ASC. Further, to relate brain synchrony and unpredictability, we correlated the GFC and CSER across regions and EEG functional bands. Exploratory analysis was not included while controlling for multiple comparisons.

## 3 Results

### 3.1 State of consciousness

The brief psychiatric rating scale (BPRS; Overall and Gorham 1962) revealed that participants were experiencing non-normal symptoms at 1 and 3 hours after intoxication, and were back to normal at 6 hours after dosing (Figure 4A., asterisks denote levels of significance). The 4B. shows that participants retrospectively reported a significant difference along the three main axes of Altered State of Consciousness scale (FDR corrected).

### 3.2 ECG: Heart rate and Heart Rate Variability

No difference between groups was observed between levels of condition psilocybin/placebo, or across time, that could not be attributed to random fluctuations in heart rate and heart rate variability. Percentage change from baseline is viewed in fig. 4 for individuals and group averages with standard error rates to account for differences between individuals.

### 3.3 EEG absolute and relative power

#### Absolute power

average increase is observed in both low and high frequencies both in psilocybin and placebo condition, with the exception of alpha and low beta frequency at 1 and 1.5 hours in the psilocybin condition, where power is below the baseline, and a broadband decrease except at alpha peak 24 hours after. Broadband increase is visible in both conditions, particularly at 6 hours. Percentage change from baseline absolute power group average is shown in 4D. Figure 5 shows that increase in delta and gamma is localised in temporal electrodes. Z-score changes from baseline per participant are in fig. 6 A., showing decrease in alpha and beta; effect size reaching significance at 1, 1.5 and 3 hours, and significant increase in delta and gamma frequencies, after temporal channels were removed. A median group difference is present in fig. 6B. where size of effect reaches significant decrease at 1.5 hours in alpha and beta at 3 hours, and shows a trend toward a decrease at 3 and 6 hours, and a broadband decrease at 24 hours (Z score of *> ±*1.96).

**Figure 5:**
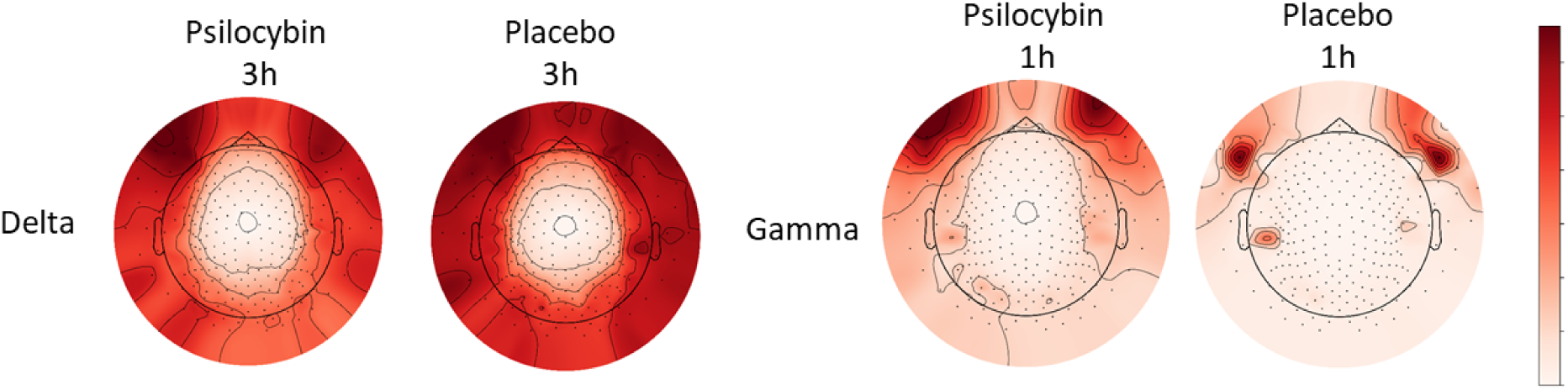
The topographical maps of median group activation in delta and gamma frequencies. The activation is from activity recorded under electrodes on the cheeks. Median is displayed as mean was affected by single electrode (outlier) activation. Temporal electrodes were excluded from further group statistical analysis (see fig. 2).

**Figure 6:**
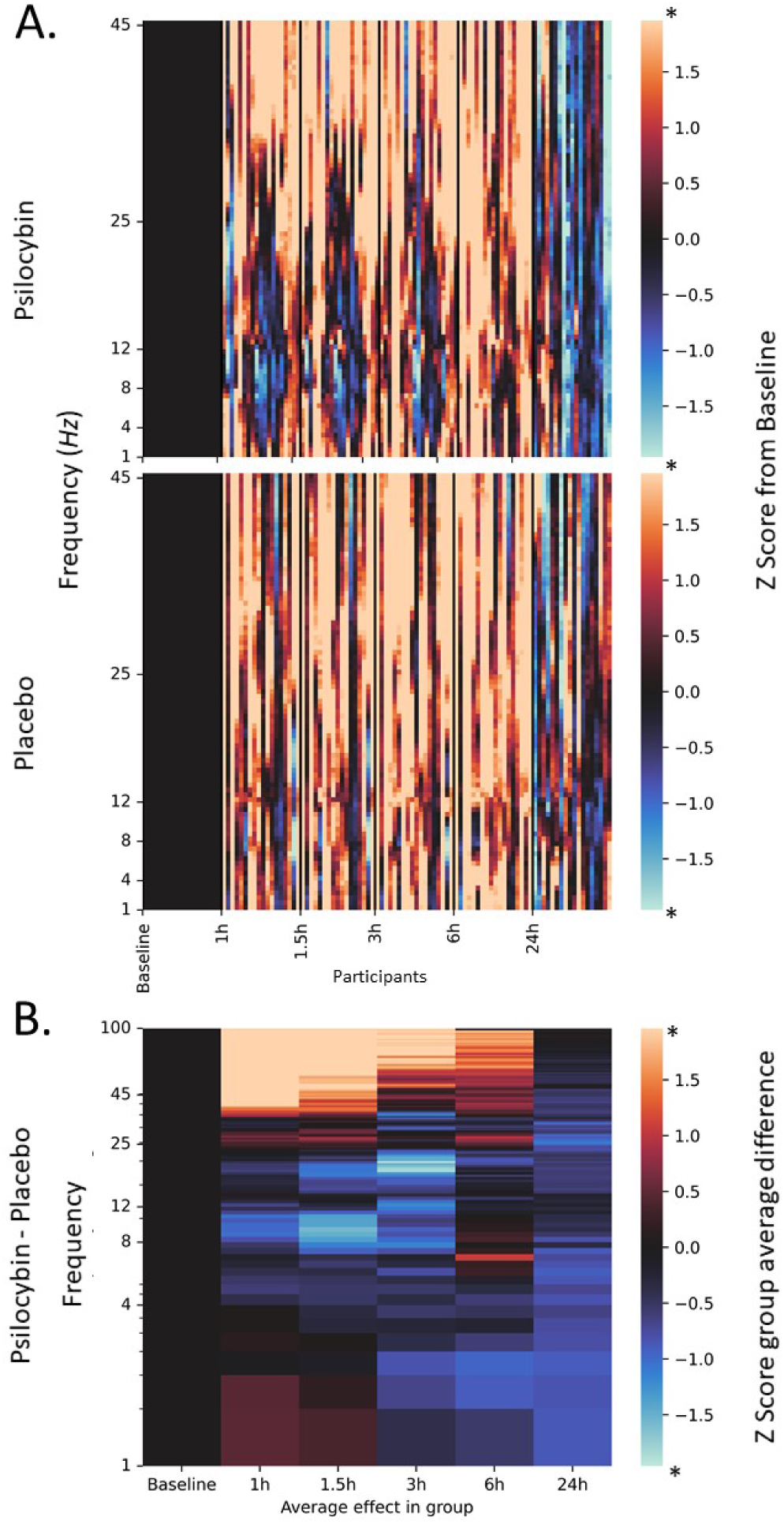
**A. Z score change from baseline in Psilocybin and Placebo.** Most prominent changes are decrease in alpha and beta activity. **B. Difference between Psilocybin and Placebo group** shows significant increase in high frequency synchronisation and trend towards a decrease in alpha and beta bands during intoxication. At 24 hours there is a trend towards a decrease in absolute power. Values further than absolute distance of 1.96 from zero are associated with *p* < 0.05.

#### Relative power

obtained for each individual representing spectral composition was compared between psilocybin and placebo groups and between functional bands (delta 1-4Hz, theta 4-7Hz, alpha 8-12Hz, beta 13-25Hz, gamma 25-45Hz) at 6 time points computing a two-tailed permutation analysis (*α* = 0.05). Temporal channels known to include a lot of muscle artifacts were excluded from the analysis (see fig. 5 and fig 2).

Individual relative power is shown in figure 7. A drop in relative power from baseline in the alpha band can be seen in two participants (PSI025 and PSI026) with return to normal at 3 hours. We observed a significant reduction in power in the psilocybin condition in the Alpha band at 1, 1.5, and 3 hours compared to placebo. At 3 hours, we also observed a significant decrease in beta activity frontally (fig. 8b, relative power, cluster analysis).

**Figure 7:**
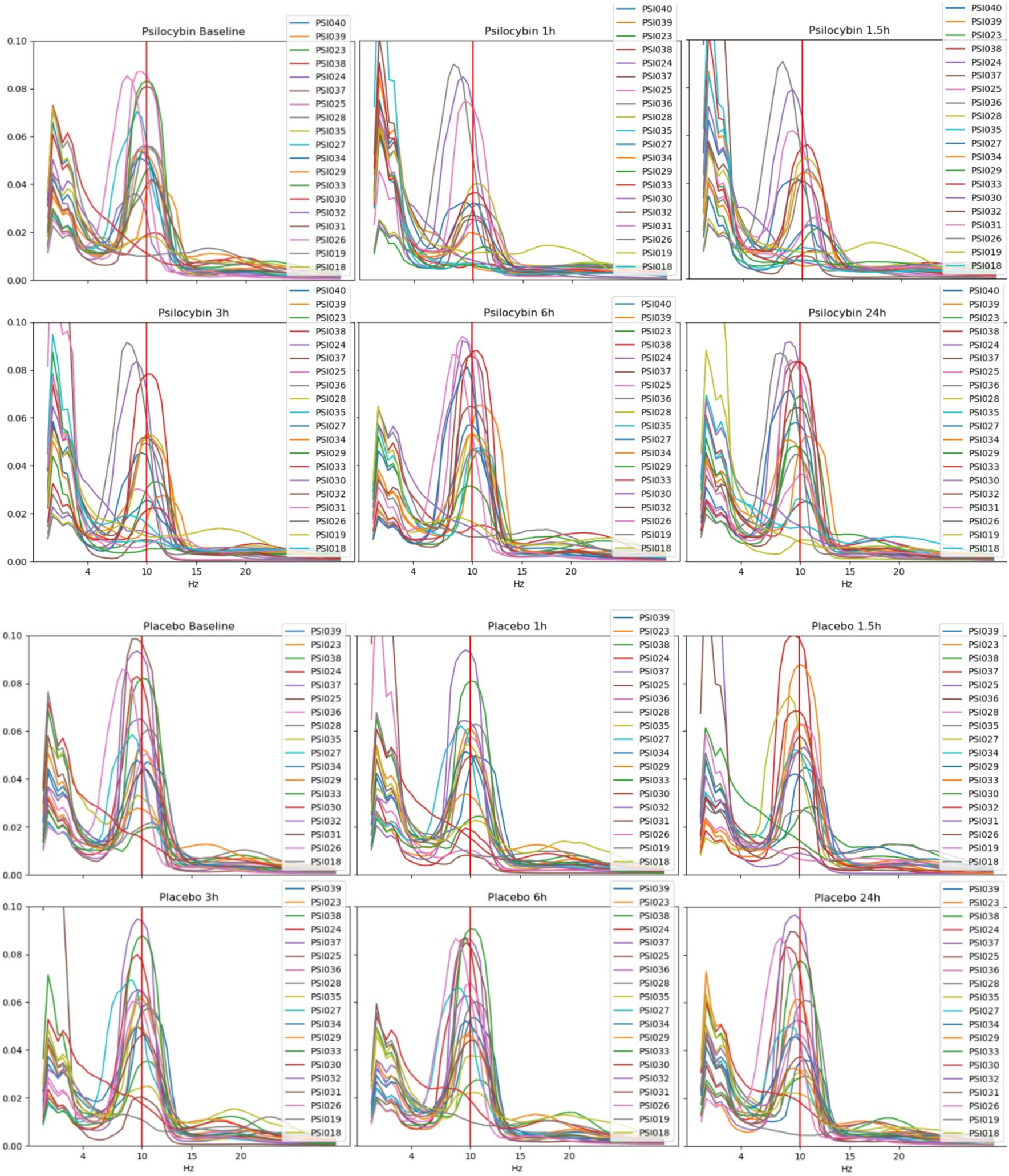
**Relative power across conditions and times** describe frequency energy composition changes for each individual.

**Figure 8:**
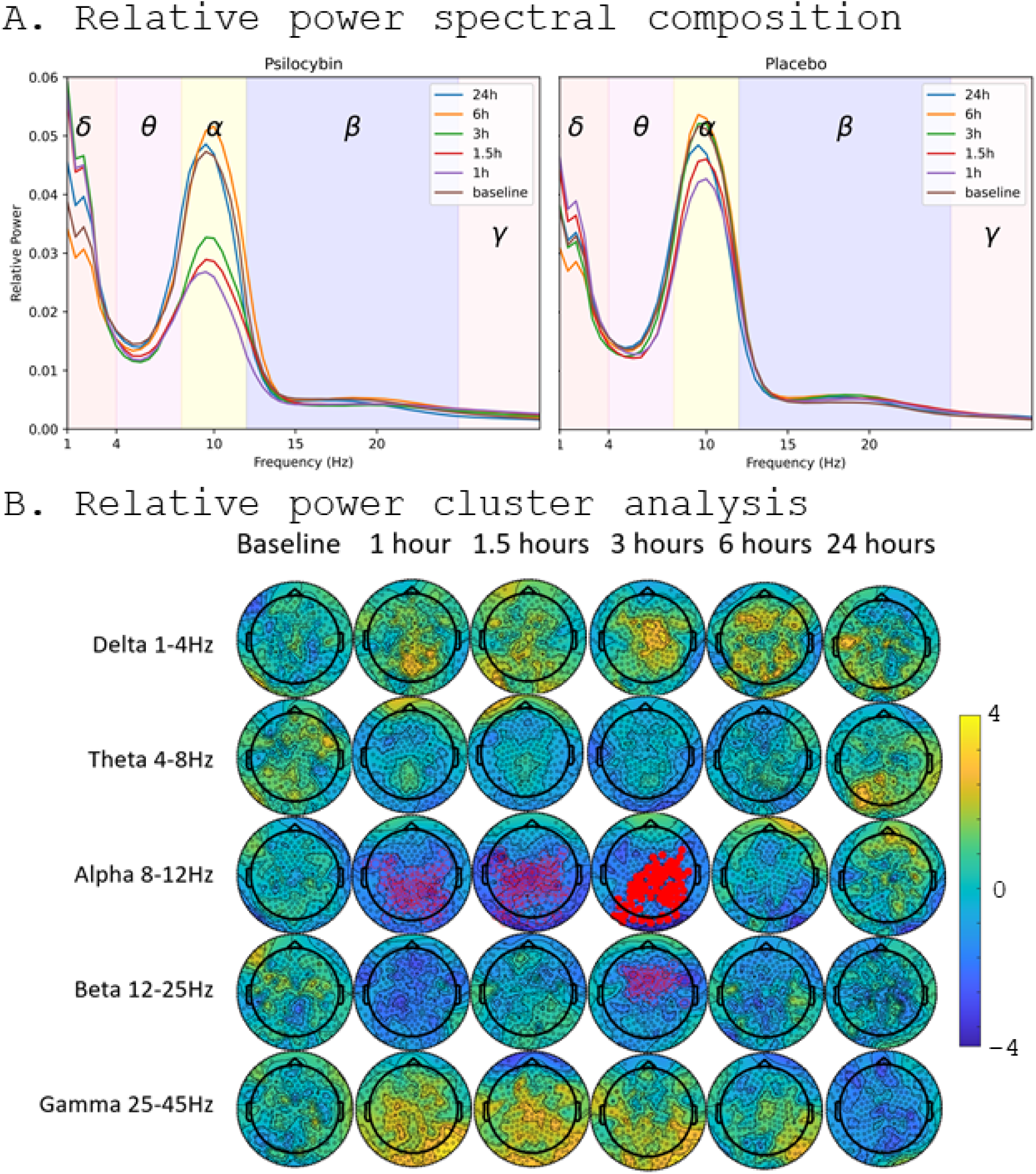
**A. Group average relative power spectral composition across time** shows drop in alpha EEG functional band (8-12Hz) at 1, 1,5, and 3 hours. **B. Relative power spectral density permutation based cluster analysis** shows changes in relative power distribution. Compared to placebo, a significant reduction frontally, parietally and occipitally occurred at 1, 1.5, and 3 hours. Reduction in Beta was observed at 3 hours. Full red mark significance at *p* <0.01 and hollow red marks at *p* <0.05. Topomaps show us a trend towards an increase in delta, and gamma bands at 1-3hours. T values shown.

**Figure 9:**
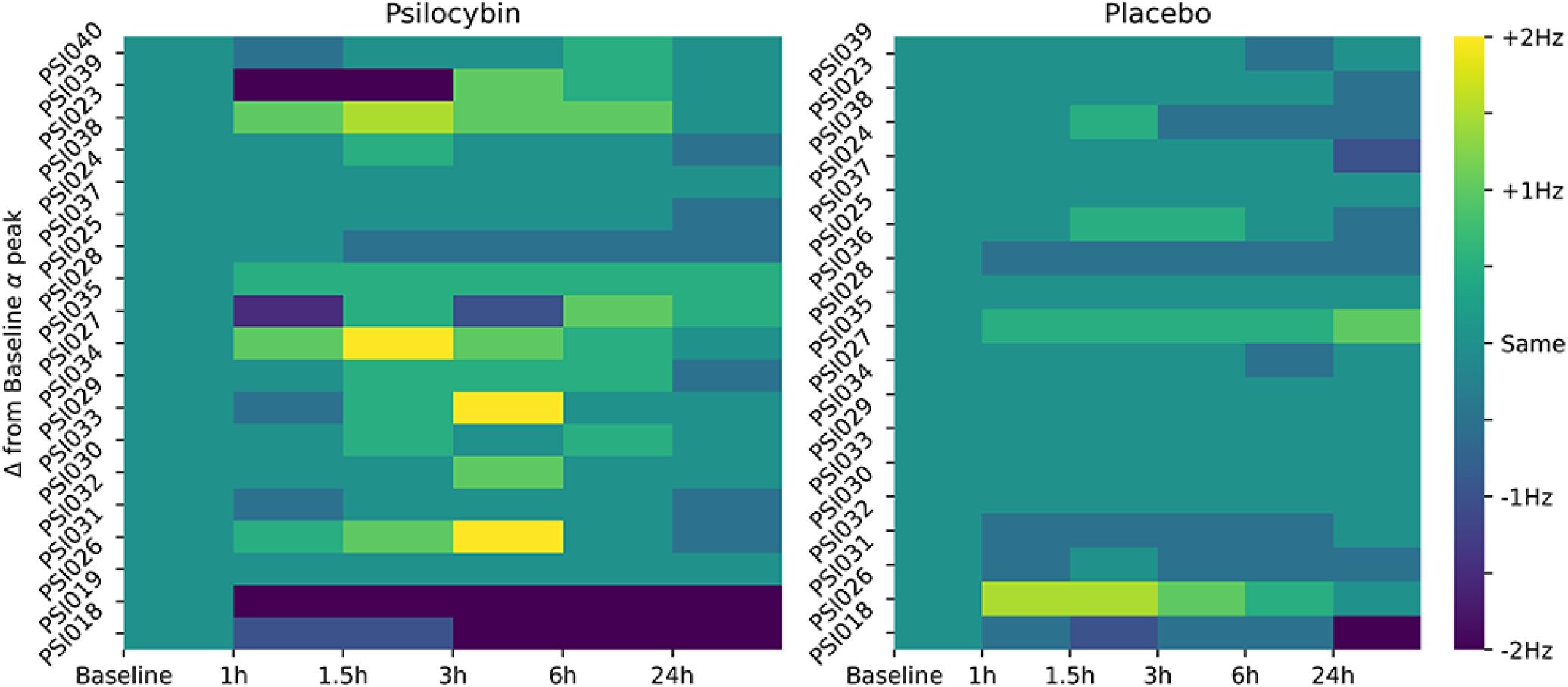
Change from baseline **Alpha peak** across conditions and times for each individual. No significant difference from baseline was observed, likely due to the opposing effects on each individual.

**Figure 10:**
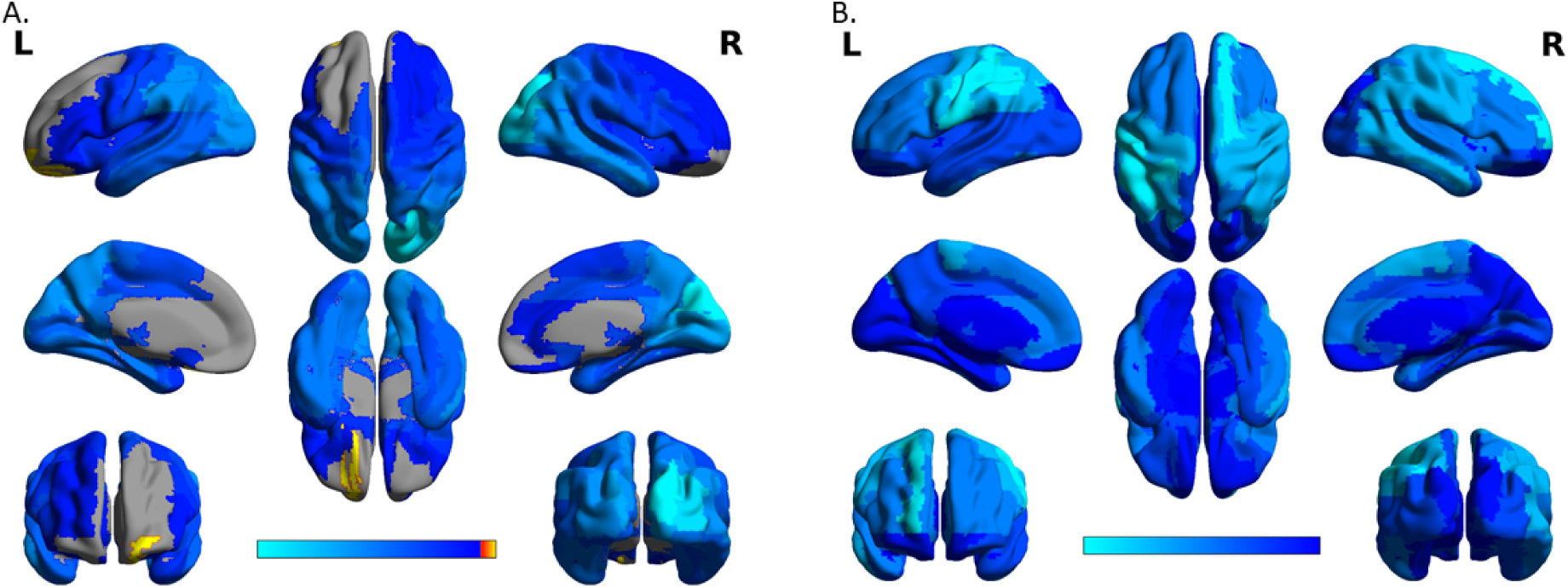
Source localisation of changes from placebo in **A. alpha band at 3 hours** post ingestion shows reduction in alpha EEG functional band (8-12Hz) activity in the right hemisphere Occipital superior and middle, Cuneus, Calcarine areas. **B. Beta band at three hours** shows decrease in activity mainly in Postcentral, Precentral, and Parietal Inferior left hemispheric areas and Frontal Superior on the right side.

### 3.4 Global Functional Connectivity

The GFC shows mainly a whole brain decrease in connectivity in the alpha band at 1 and 1.5 hours, and a decrease parietally in the beta frequency at 1 hour in psilocybin group. In placebo, an increase in delta band at 6 hours frontally and at 1.5 hours parietally is observed. Inverted individual difference from baseline, and group averages are shown in fig. 11 with error bars and asterisks marking FDR corrected significant difference obtained from an LMM model. Fig. 16A. shows GFC averaged over the source reconstructed space, baseline removed. Significant results of the LMM model did not survive correction. A trend towards a decrease was observed in alpha at 1 and 1.5 hours globally, and an increase in the DMN in the gamma band at 1 hour. Note that while significant group differences were observed in the alpha, beta, and gamma band, the opposite effect on individual level is also visible. Further, in the placebo condition, changes from baseline of similar magnitude can occur.

**Figure 11:**
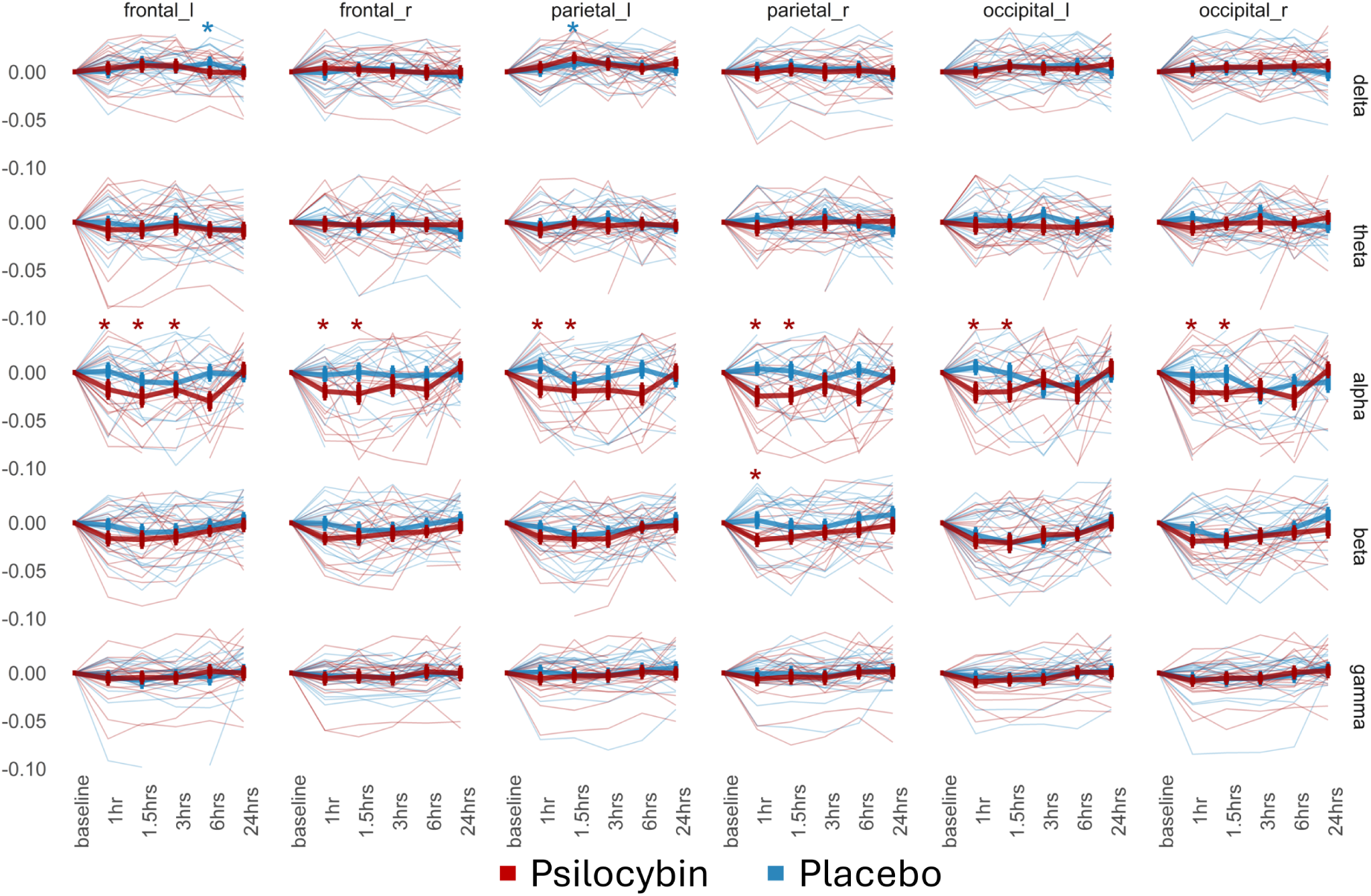
Change from baseline in **Power Envelope GFC** across functional frequencies averaged over frontal, parietal, and occipital regions laterally shows significant decrease from baseline in connectivity globally at 1 and 1.5 hours, and frontally also at 3 hours in the psilocybin group. On the right parietal lobe we also see decrease in beta at 1 hour, and in the placebo group we see an increase in delta activity.

**Figure 12:**
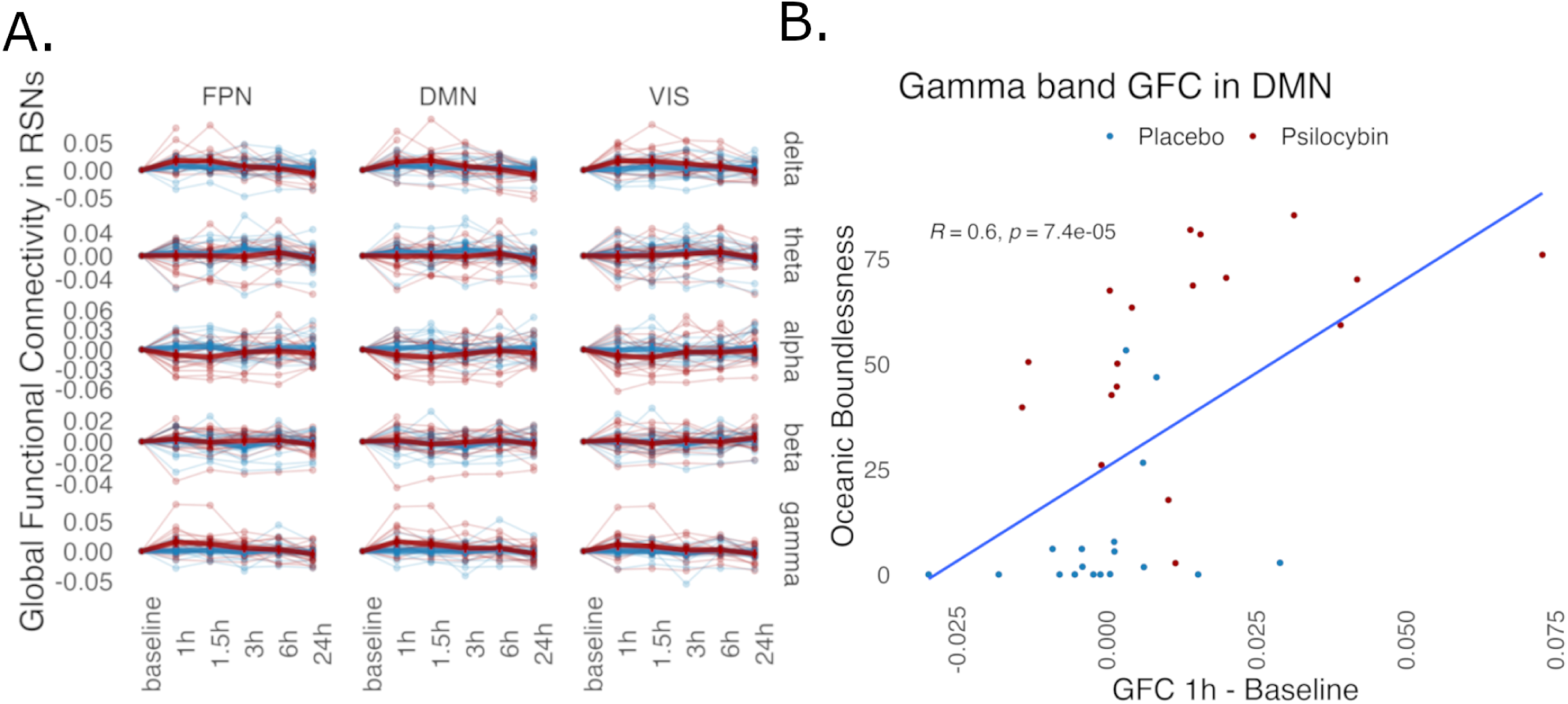
**A.**Change from baseline in **GFC RSNs** across functional frequencies. Comparisons did not survive FDR correction. We see a trend towards a decrease in all networks in alpha band at 1 & 1.5 hours, and an increase in gamma in DMN at 1h. **B. Change in GFC from Baseline in DMN gamma band** shows a moderate Pearson correlation between magnitude of change, and intensity of oceanic Boundlessness subscale of the altered states of consciousness.

**Figure 13:**
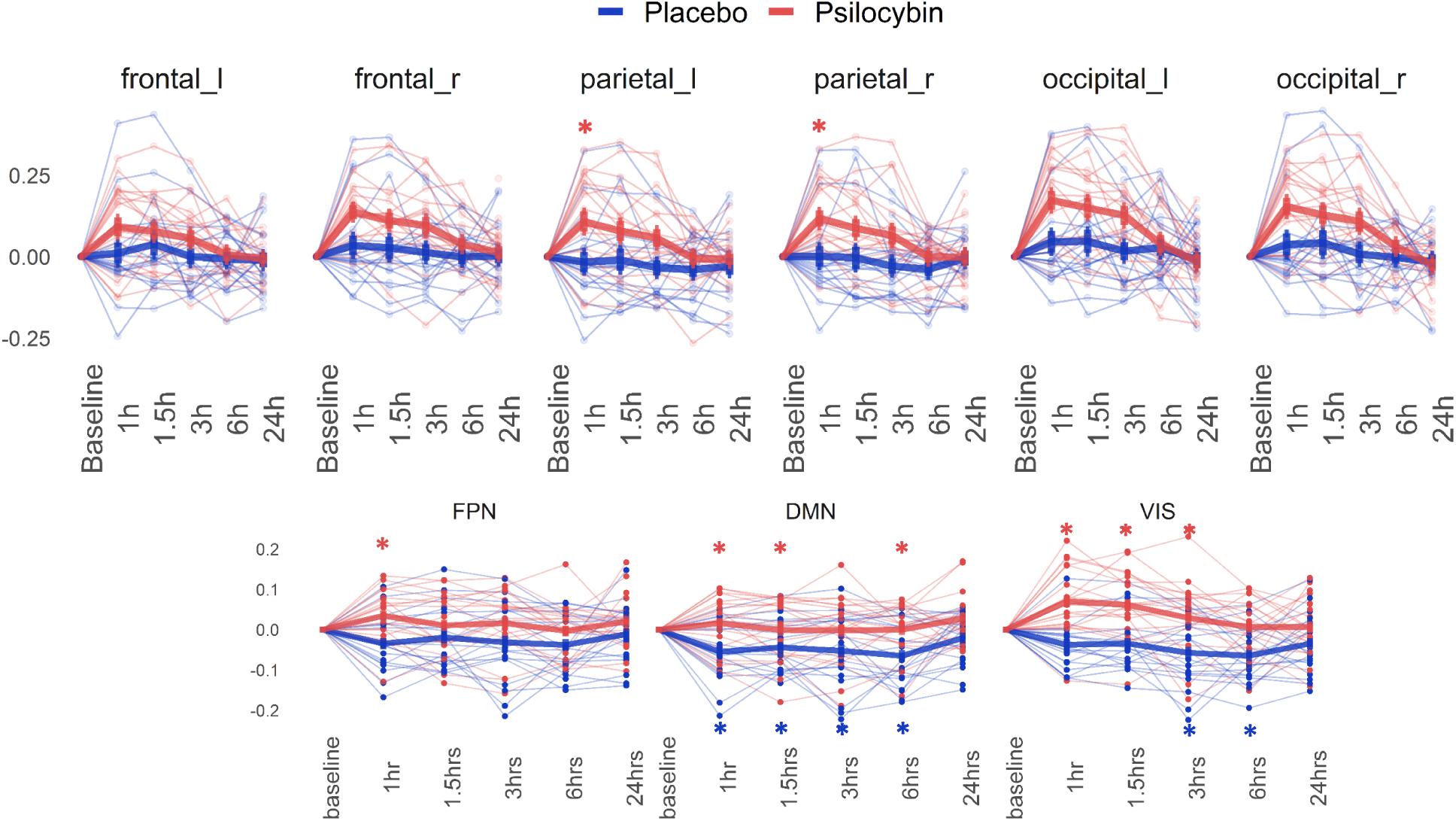
Change from baseline **Lempel-Ziv Complexity** normalised entropy rate shows rapid increase in entropy at 1 hour parietally. In the source-space, increase in entropy is observed in the psilocybin group at 1 hour in the frontoparietal network, at 1, 1.5, and 6 hours in the Default Mode Network, and 1, 1.5, and 3 hours in the Visual Network, and 1, and 3 hours in the rest of the brain. In the placebo condition, a drop in complexity is seen from 1 to 6 hours in the default mode network, and 3 and 6 hours in the visual network.

### 3.5 Entropy measures

#### 3.5.1 Lempel Ziv Complexity

LZ signal diversity showed increase at 1 hour in the parietal lobes. When averaged over the 3 functional networks in the AAL90 space, significant increase in entropy is observed in the psilocybin condition, mainly in the default mode network and the visual network. A significant decrease in entropy is observed for the duration of the experiment in the placebo condition, particularly in the default mode network, see fig. 13.

#### 3.5.2 Complexity via State Space Entropy Rate

Psilocybin reduced CSER complexity in the alpha band globally at 1 to 3 hours, and an increased broadband complexity at 1h parietally and in the right frontal lobe. In the placebo condition, a decrease was observed in the delta band (fig 14). Analysis over functional networks in the source-space revealed that complexity decreased in the alpha band in the visual network and rest of the brain at 1 to 3 hours, in the frontoparietal network at 1 and 1.5 hours, and at 1 hour in the DMN. A significant increase in complexity from 1 to 6 hours is observed in the gamma band in the visual cortex. In the placebo group, an increase in complexity is seen in the placebo group in the FPN cortex in alpha and beta, and a decrease in the DMN at 1, 3, and 6 hours broadband. Interestingly, in the visual cortex, there is a significant increase at 3 & 6 hours in the alpha, but a decrease in gamma (15).

**Figure 14:**
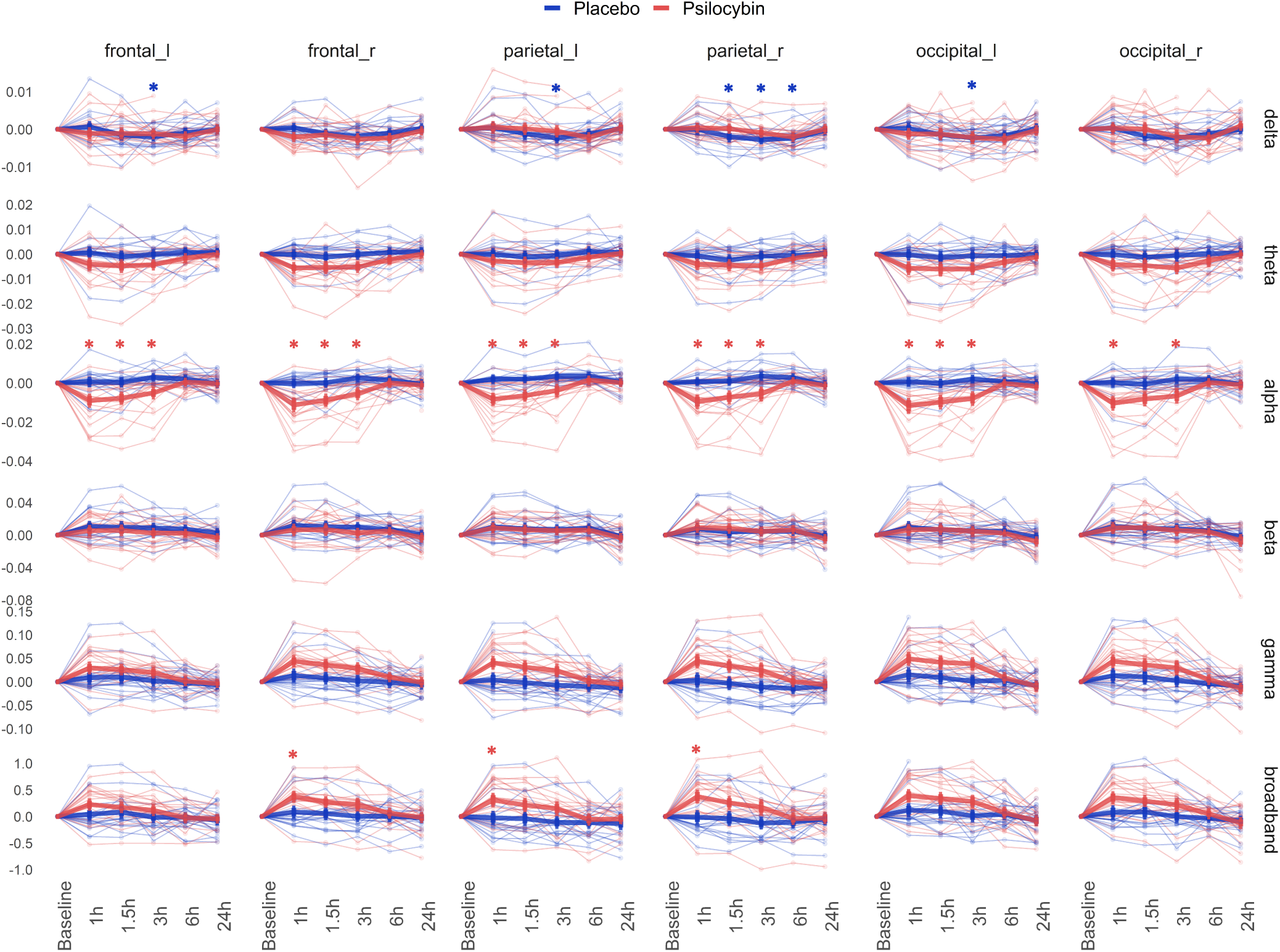
Change from baseline **Complexity via State Space Entropy Rate** shows a distinct change in psilocybin condition a decrease in alpha band at 1, 1.5, and 3 hours globally, and an increase at 1 hour parietally, and frontally on the right hemisphere. Broadband CSER shows similar results to LZ, and an opposing effects of psilocybin on alpha and broadband entropy, and further that entropy is mostly driven by higher frequencies. A decrease in entropy in the placebo condition is observed parietally and on the frontal and occipital left hemisphere. Note different scales on the Y axis.

#### 3.5.3 EEG measures in the DMN and subjective experience

Changes in default mode network from baseline were explored. Only the change from baseline in gamma GFC in DMN at 1 hour showed a moderate significant relationship between intensity of subjective experience represented by the Oceanic Boundlessness. Due to low variability and the floor effect in the placebo condition, this needs to be interpreted with caution. We also correlated changes in other functional bands in the DMN with OBN, but these were not significant and we do not report them.

#### 3.5.4 GFC and CSER

Fig 16 shows the relationship between absolute values of CSER and GFC at different time points and across functional bands, with Pearson correlation and associated p-value, uncorrected. Note that the x scale differs for each EEG band.

## 4 Discussion

In a cross-over design, we compared placebo with 0.26mg/kg of psilocybin administered orally (20mg in 77kg person), a low to moderate dose. Healthy subjects experienced significant alteration to the normal state of consciousness on sessions when they received psilocybin compared to placebo. Changes started within 1 hour, and were back to normal levels at 6 hours (fig 4 A. and B.). These alterations did not result in significant changes to the heart rate and heart rate variability (fig 4 C.) but resulted in changes in EEG activity (fig 4 D.). In accord, Holze et al., 2022 shows that low dose of psilocybin (15mg) lasted app. 4 hours, whereas high dose (30mg) lasted app. 7 hours, that the low dose did not cause changes in the heart rate but the high dose did.

Absolute EEG power increased in high (gamma) and decreased in alpha and beta frequencies compared to placebo during the session, and 24h after dosing there is a trend towards a decrease broadband (see figs. 4D., and 6). Changes in delta and gamma were induced by activation in peripheral electrodes (fig. 5) likely originates from jaw clenching rather than from neural activity, and temporal channels were removed from further analysis. An increase in low and high frequencies was present in both placebo and psilocybin conditions, and may be associated with setting of EEG gel, and galvanic skin response to the warm conditions inside the Faraday cage. Broadband power desynchronisation after psilocybin was observed previously in rodents (Tyls et al., 2016), and in humans with psychedelics in general (Le et al., 2024).

Analysis of relative power composition showed that psilocybin administration resulted in attenuation in the alpha band at 1 hour, 1.5 hours, and 3 hours globally and an attenuation in beta band frontally (fig 7 and 8). Source reconstruction localised sources of the decrease at 3 hours in alpha band mainly on the right hemisphere in the occipital superior and middle, and the cuneus and calcarine areas. In the beta band the decrease was mostly in the postcentral, precentral, and parietal inferior areas on the left hemisphere, and in the frontal superior right area (fig. 10). Attenuation in the alpha band activity is a robust trait of a psychedelic state associated with visual experience of imagery, seeing with eyes-closed (R. L. Carhart-Harris et al., 2016; De Araujo et al., 2012), which may be caused by deficient thalamic gating in the pulvinar (Preller et al., 2019) affecting activity in the V1. We did not observe a shift in the alpha peak under psilocybin, our data shows that some individuals experience quickening and some slowing of the alpha activity (fig 9). We observe a significant decrease in beta spectra at 3 hours (fig. 8). It is intriguing that this effect is temporarily localised to three hours, even though a trend towards this is observable at 1 hour as well. The desynchronisation of the frontal beta activity may generally be associated with inability to maintain focus, perform cognitively demanding tasks, and from inability to suppress thoughts (Bonnieux et al., 2023; R. L. Carhart-Harris & Friston, 2019). From our data we may conclude that psilocybin becomes active first in the occipital regions, then in the frontal regions, experienced as intense visual stimulation only later followed by change in cognition.

The global functional connectivity (GFC) revealed a significant decrease in connectivity from baseline globally in the alpha band at 1 and 1.5 hours, and at 3 hours frontally (fig 11), and a decrease in beta GFC at 1 hour parietally. Comparing groups on GFC across RSNs did not survive FDR correction, however a trend towards a decrease in alpha band and an increase in gamma band is visible (fig. 12 in the DMN). It has been shown that psilocybin produces broadband desynchronisation of activity in MEG signal (Muthukumaraswamy et al., 2013). Siegel et al., 2023 report similar findings from fMRI blood oxygen level dependent (BOLD) signal, that the decrease is mainly in the Default Mode Network. R. L. Carhart-Harris et al., 2013 show that the functional connectivity from DMN is however increased to task positive networks as a result of psilocybin, a relationship which is orthogonal otherwise. Decrease in GFC may reflect two things. Firstly, decrease in information processing overall, or, secondly, increase in information processing locally. Because power increased, but GFC decreased, it may be deduced that information processing did not cease, but became segregated as a function of psilocybin. Decrease in GFC is associated with cognitive decline (e.g. (Onoda et al., 2012)), but coupled with known effects of psilocybin, it may reflect impairment in cognitive functioning perhaps due to over-integration of over-salient stimuli. In other words, inability to focus by being distractable by external and internal processes (R. L. Carhart-Harris et al., 2011).

If stimulus integration into neural activity is segregated, i.e. processed locally, then the brain activity would reach more uncoordinated neural states, which would increase the complexity of the EEG traces. Analysis of signal diversity using the Lempel-Ziv (Lempel & Ziv, 1976) algorithm shows an increase at 1 hour parietally. However, a significant increase in LZ is observed in the visual network at 1 to 3 hours, in the DMN at 1, 1.5, and 6 hours, and in the frontoparietal network at 1 hour. Increase in LZ was reported in LSD condition (Mediano et al., 2024), DMT (Timmermann et al., 2023) and Psilocybin (M. M. Schartner et al., 2017). But an increase in LZ is also associated with changes in information processing due to acupuncture (Luo et al., 2012) and after processing complex visual and auditory stimuli (Orłowski & Bola, 2023). Thus, LZ may increase when the brain is trying to interpret a not-easily interpretable stimulus. When psychedelics increase brain segregation, the brain struggles to process information locally. This may be represented psychologically by experiences of novelty. Interestingly, we also observe a decrease in signal diversity in the placebo condition mainly in the default mode network from 1 to 6 hours, and the visual network at 3 and 6 hours (fig 13). From perspective of stimulus integration, this may be explained as a habituation to the study conditions. Changes in the visual area thus may be associated with vivid imagery, whereas changes to DMN may reflect perception of self; a process which may be heightened in the psilocybin condition, but relaxes over time under in placebo condition.

Complexity via State-Space Entropy Rate results show that signal diversity decreases globally under psilocybin in the alpha band at 1 to 3 hours, and increases in the higher frequencies at 1 hour (see fig. 14), which replicates the findings of Mediano et al., 2023. CSER analysis over RSNs shows decrease mainly in the alpha band in the visual network at 1 to 3 hours, and the fronto-parietal network at 1 and 1.5hours, and an increase in the gamma band at 1 to 6 hours in the visual network, but also a sporadic increase in DMN. The visual cortex shows sensitivity to changes induced by psilocybin as measured by decrease in CSER and LZ. An apparent relationship between alpha decrease and higher frequencies increase in the psilocybin condition may reflect integration of stimuli behind closed eyes. In a normal wake eyes-closed resting state, alpha activity in the visual cortex may be interpreted as synchronisation of weakly coupled oscillators receiving random input (Hidalgo et al., 2022). Complexity may be driven down when the signal becomes non-random due to changes induced by psilocybin, marked by desynchronisation in the alpha power spectra. However, associative processing of drug induced content may be happening in higher frequencies gamma. Our findings make case for the careful study of higher brain frequencies, a task difficult due to EEG artifacts. In the placebo condition, we see a significant increase over the fronto parietal network from 1 to 6 hours in both alpha and beta bands, but also decreases in the higher frequencies over the default mode network and the visual area; we see the opposite effect to psilocybin condition, that CSER increases in alpha at 3 & 6 hours and decreases in higher frequencies (fig 15). These changes are intriguing, they may reflect on processes natural to prolonged relaxation, in any case different to psychedelic experience. Changes in band-specific complexity need to be studied further to allow for interpretation.

**Figure 15:**
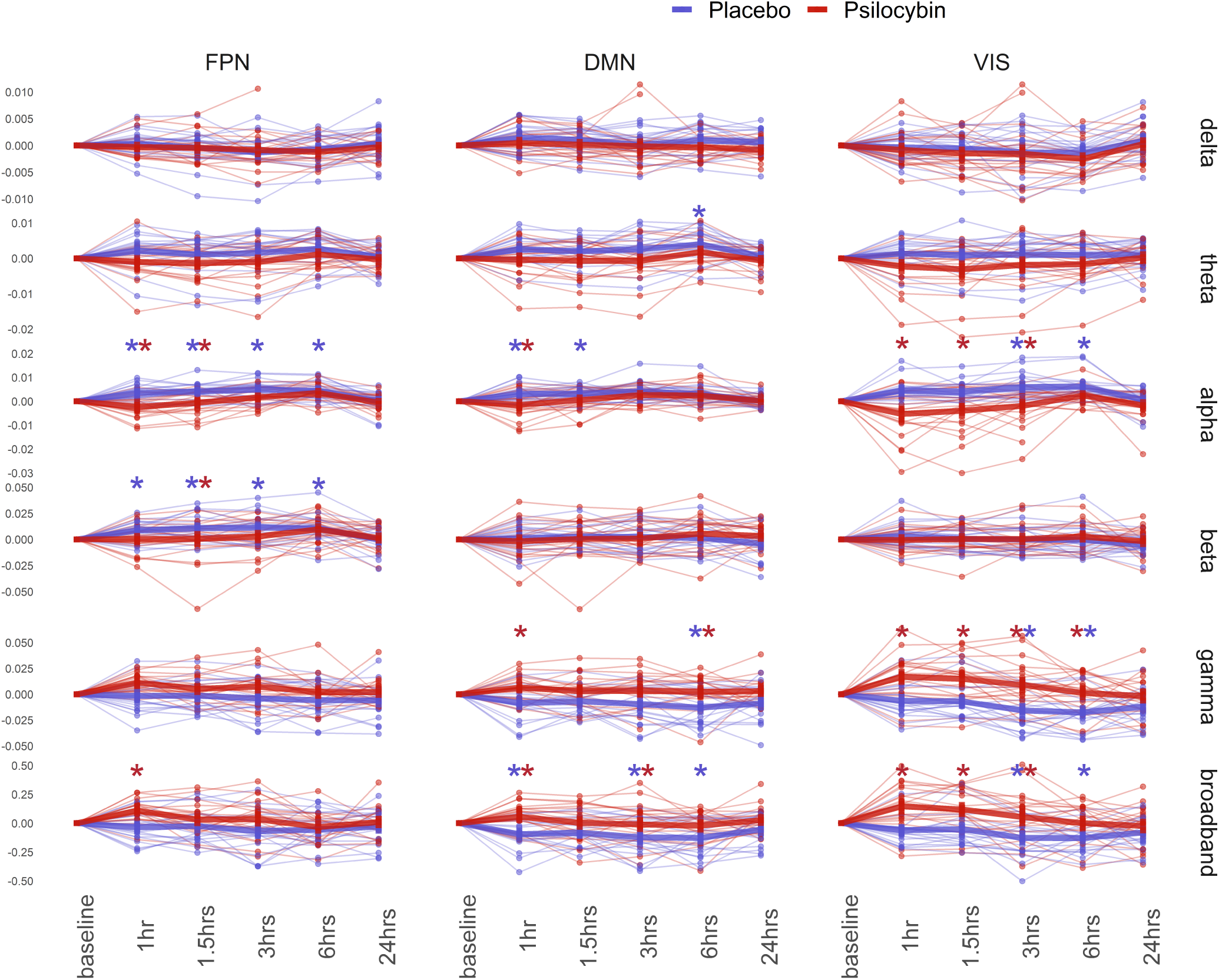
Change from baseline **Complexity via State Space Entropy Rate** normalised entropy rate over source-space functional networks shows a decrease in entropy mainly over the visual network at 1-3 hours in the alpha band, and an increase in gamma and higher frequencies. Interestingly, we can observe an increase in CSER in the alpha and beta band in the frontoparietal network, and a decrease in the default mode network and visual network in the broadband.

**Figure 16:**
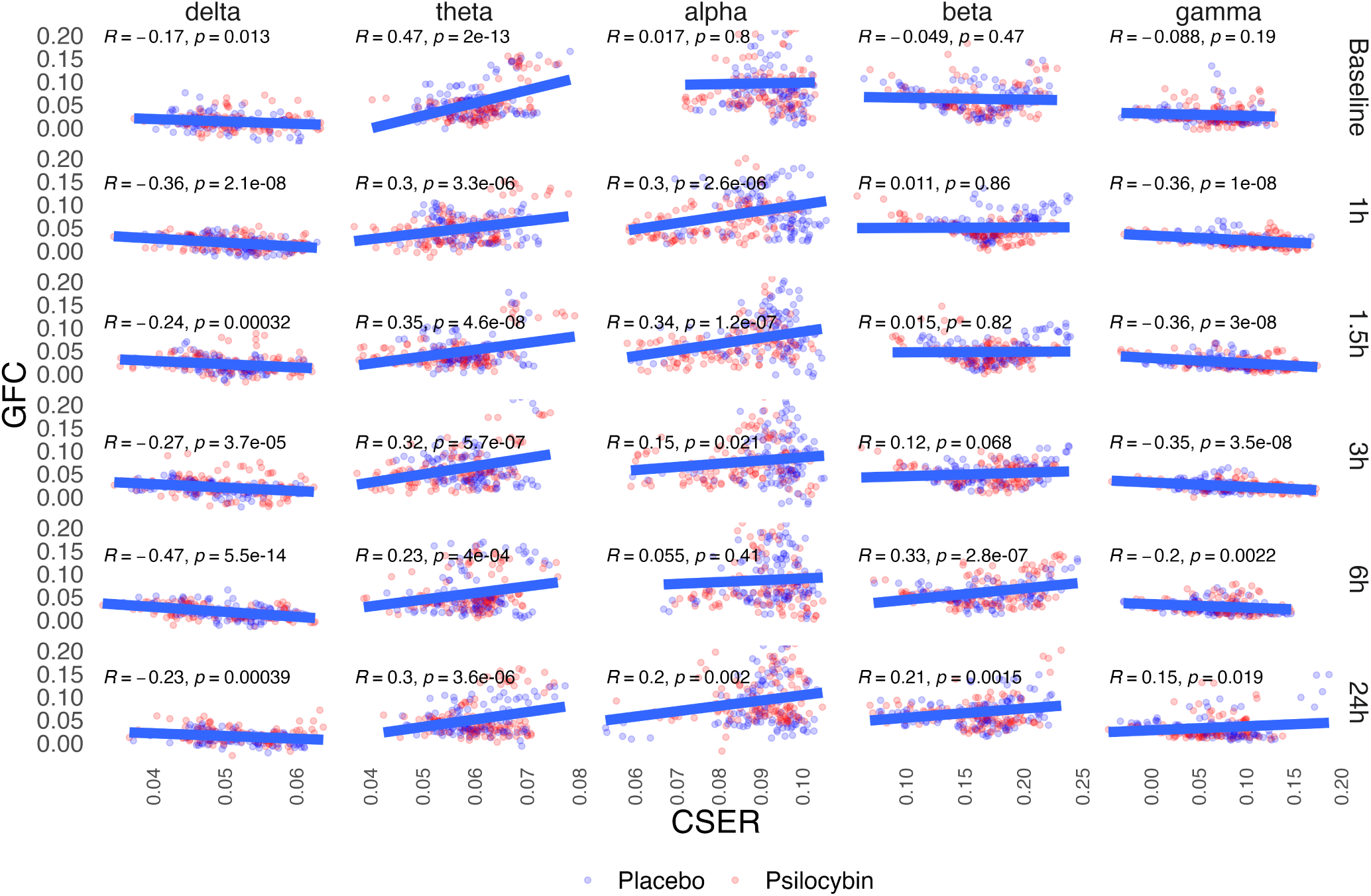
Functional band decomposition **correlation between GFC and CSER**. From left, weak negative relationship appears to be in delta, weak positive relationship in theta, but in alpha a weak positive relationship arises during the 1-3 hours, suggesting that psilocybin alters this relationship, and an opposite relationship is present in gamma. In beta, a weak positive relationship appears at 6 and 24 hours. Pearson correlation calculated, uncorrected. Lines represent general linear models.

We found a relationship between a change from baseline to 1-hour in GFC in gamma band in the DMN, and subjective experience as captured by Oceanic Boundlessness sub scale of the ASC questionnaire, but not for LZ or CSER. Our exploratory finding suggests that the more DMN becomes connected with the rest of the brain, the more individuals experience a sense of unity. The DMN, active during passive rest and mind wandering and connected to self-reference, becomes under psilocybin more functionally connected with the rest of the brain, which may explain the intensity of the psychedelic experience, perhaps as the self is integrated with the rest of the brain, thus effectively lowering the segregation of both our sensory information, associations about the world, and self. When correlating the GFC and CSER values, we find an emergent relationship during the hours of effect of psilocybin in alpha and gamma bands. This emergent relationship shows that the lower the GFC, the lower the CSER in alpha, and conversely the lower the GFC, the higher the CSER in gamma. If alpha activity is connected mostly to processing of the visual stimuli, and gamma mostly with the higher order associations, then the intense visual imagery during psilocybin are associated with less unpredictable signal, i.e. visual experience is processed locally. However, the gamma has the opposite relationship, gamma signalling under psilocybin is processed locally, more segregated and more unpredictable. Our evidence may be viewed in support of the REBUS theory that psychedelics work to relax high-level priors on both sensory and associative level (R. L. Carhart-Harris & Friston, 2019), which leads to both segregation in the sensory cortex as marked by decrease in power and GFC in the alpha band, but also an integration of the DMN as the high-level priors are being re-addressed by propagation of bottom-up prediction-errors, as marked by the increase in gamma signal complexity.

In summary, we found difference between the psilocybin and placebo group on measures of subjective intensity of experience (fig. 4), and in EEG measures of Power, GFC, LZ, and CSER (see 6, 8, 11, 13, 15), but we observed a linear relationship only between DMN gamma GFC and subjective experience (fig. 12). Two possible explanations arise, A. changes in the brain do not relate to psychological intensity of experience, or B. intensity of psychological experience is not only related to dose-dependent neurobiological changes captured by EEG, but also dependent on a complex pattern of personal experience including immediate mood and long-term traits (Studerus et al., 2012), generally referred to as Set in Set and Setting (e.g. Hartogsohn, 2016). Our findings are complementary to emerging evidence that neuroimaging changes correlate with symptoms improvement in depression patients (Kuburi et al., 2022), with changes in brain activity mainly in the DMN, however, these changes are insofar too insensitive to use reliably as a predictor of treatment outcome.

## 5 Limitations

This study was part of a clinical trial. Dosage was weight dependent, which is currently not the standard. Dose was administered orally, which may lead to different metabolisation speed, biasing the effects of psilocybin at any given time point in a small sample. EEG measures used are sensitive to noise in the signal. Techniques beyond removal of obviously affected epochs are limited. Is not the topic of this work to prove that EEG gamma activity is of neural origin. We show that changes in the gamma range differ across the psilocybin/placebo conditions, and that these changes should be further studied in relation to psychedelics. Thus interpretation of our results should be cautious. The within subject design has limited validity in an EEG study with variable length between dosages, as the EEG traces may change in time in themselves and together with dependency on electrode position and impedance. Functional unblinding appears problematic in all studies with psychedelics. In our case, if participants experienced psychedelic-like state on their first visit, they would be unblinded to the second visit. Perhaps partly because of the reasons above, we observe a variation in our measures marked by that *≈* 10 % of participants in either group show results similar to average of the opposite group; in EEG measures and the subjective measures of intensity of experience measured by the ASC. This fosters the notion that the role of the set and setting in which such experience is meant to take place, is crucial.

## 6 Conclusion

Psilocybin affects EEG activity acutely. The dominant change is the decrease in the alpha and an increase in the gamma power, followed by a decrease in beta. This decrease, a local desynchronisation, is marked by the decrease in global functional connectivity. Signal complexity increased, particularly in the visual area and DMN. Decomposition of complexity into functional bands shows decrease in complexity in the alpha band but an increase in the gamma band. These changes are time dependent, and are likely related with the relaxation of strong believes about the world, and integration of self with the environment as marked by the experience of unity. We could not confirm associations between baseline values or changes in magnitude of EEG measures as intensity of the psilocybin experience, except for changes in GFC in DMN in the gamma band.

## Data Availability

The data that support the findings are available on request from the corresponding author. The data are not publicly available due to privacy or ethical restrictions.

## Author Contributions

TP run the study as a principal investigator. TF guided the analysis of signal complexity. MN carried out the current analysis. PM assisted with computational approaches. All authors contributed to the manuscript.

## 7 Acknowledgements

This work was supported by the Czech Science Foundation (projects 23-07578K), Czech Health Research Council (project NU21-04-00307), the national budget through MEYS, LRI CZECRIN (LM2023049), Long-term Conceptual Development of Research Organization (RVO 00023752), and Specific University Research, Czech Ministry of Education, Youth and Sports (project 260533/SVV/2024), the Charles University research program Cooperatio-Neurosciences, ERDF-Project Brain dynamics, No. CZ.02.01.01/00/22_008/0 Japan Science and Technology Agency via grant number JPMJPF2205, and PSYRES –Psychedelic Research Foundation. The funding sources were not involved in the study design; in the collection, analysis, and interpretation of data; in the writing of the report; nor in the decision to submit the article for publication.

## A Appendix

## A.1 Appendix 1: Inclusion and Exclusion Criteria

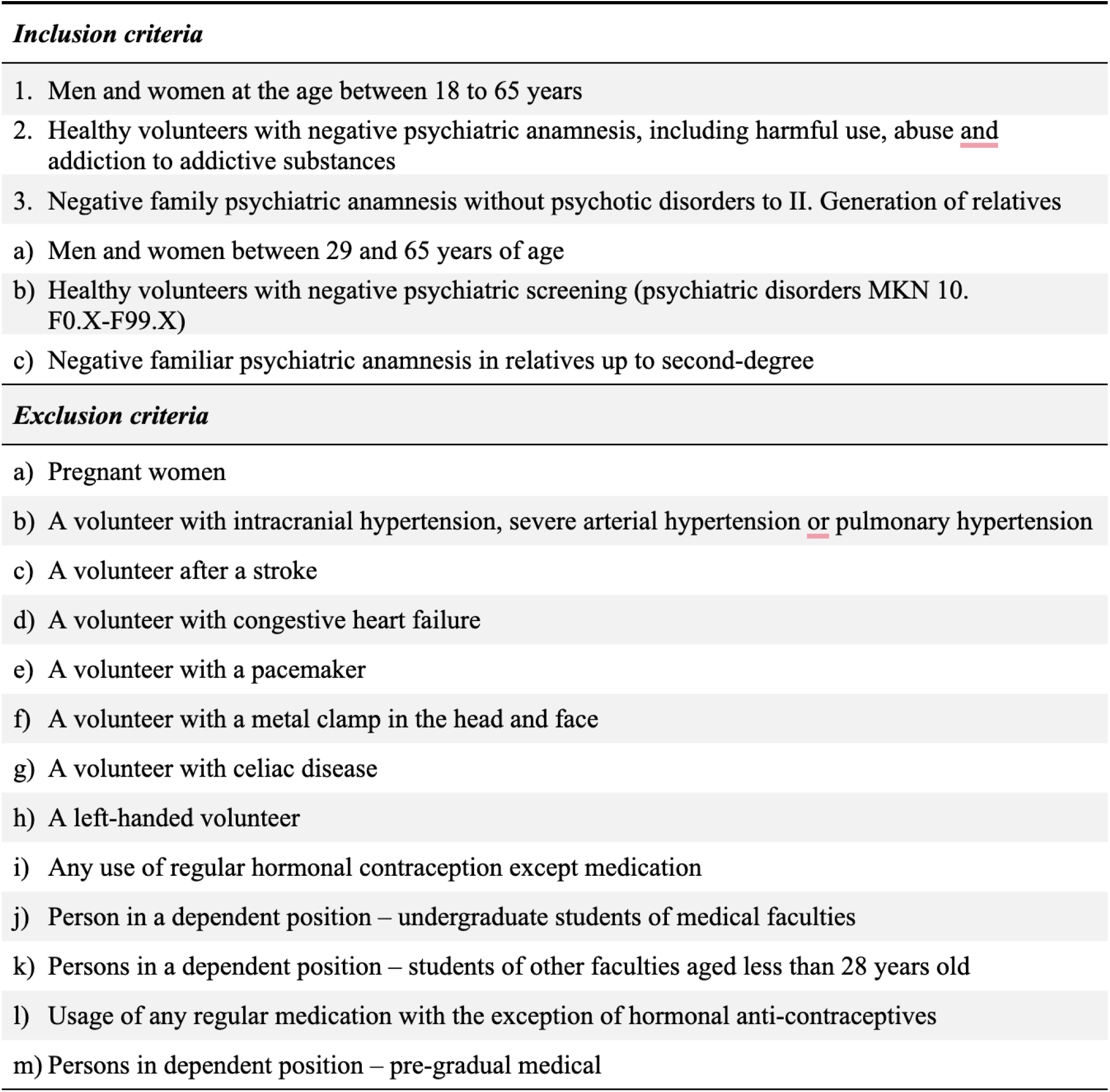

## Supplementary Material

Supplementary Material (created during production as a web link to online material).

